# A meta-analysis of genetic variant pathogenicity and sex differences in *UBQLN2*-linked amyotrophic lateral sclerosis and frontotemporal dementia

**DOI:** 10.1101/2024.10.25.24316165

**Authors:** Kyrah M. Thumbadoo, Laura R. Nementzik, Molly E.V. Swanson, Birger V. Dieriks, Michael Dragunow, Richard L. M. Faull, Maurice A. Curtis, Ian P. Blair, Garth A. Nicholson, Kelly L. Williams, Emma L. Scotter

## Abstract

Ubiquilin 2, encoded by the X-linked *UBQLN2* gene, is a ubiquitin-binding quality control protein. Pathogenic *UBQLN2* genetic variants cause X-linked dominant amyotrophic lateral sclerosis and/or frontotemporal dementia (ALS/FTD), however, clinical phenotypes from these variants show striking inter- and intra-familial heterogeneity. Further, there are many *UBQLN2* variants whose significance to disease is uncertain. Here, we examine the pathogenic potential of *UBQLN2* variants reported in individuals with ALS/FTD and their non-symptomatic relatives. Meta-analysis from 27 published studies identified 186 affected individuals and 51 asymptomatic carriers, each harbouring one of 43 unique *UBQLN2* coding variants. Features of identified variants, including evolutionary conservation, minor allele frequencies, localisation to protein domains, and *in silico* predictions of pathogenicity were compiled. Per biological sex, clinical features were compared between *UBQLN2* variants segregated by pathogenicity. Pathogenic *UBQLN2* variants carriers, of which most are familial ALS cases, showed a sex-specific difference in age at onset wherein males developed disease on average 18.15 years prior to females (29.54 ± 11.9 versus 47.69 ± 13.4 years; p<0.0001), with no change in disease duration (p=0.6400). *UBQLN2* variants of uncertain significance showed a bimodal distribution of onset age per sex suggesting a mixture of true benign and true pathogenic variants. In human brain tissue, two male *UBQLN2* p.Thr487Ile (ALS-FTD and ALS) cases showed a greater burden of ubiquilin 2 aggregates than a related female case (ALS-FTD). These robust sex-specific differences in ALS/FTD presentation in carriers of pathogenic *UBQLN2* variants may improve predictions of ALS/FTD risk in carriers, aiding in diagnosis and disease management.

## Introduction

Pathogenic variants in the X-linked gene *UBQLN2* cause amyotrophic lateral sclerosis (ALS) and/ or frontotemporal dementia (FTD), characterised by aggregation in the CNS of mutant ubiquilin 2 protein. ALS is characterised by the progressive loss of motor function due to the degeneration of cortical, bulbar, and spinal motor neurons^1^. FTD results from neuronal degeneration within the frontal and temporal cortices^2^. While the aggregation patterns of pathogenic ubiquilin 2 in post-mortem CNS tissues are distinct from those of wildtype ubiquilin 2^3^, many *UBQLN2* genetic variants reported in the literature are of uncertain significance and do not have patient tissue available to identify pathogenic ubiquilin 2. Yet, establishing which genetic variants within *UBQLN2* - a definitive pathogenic gene in ALS/FTD - are disease-causing is critical to understanding the aetiology of ALS/FTD and anticipating clinical impact.

While traditionally seen as a motor disorder, up to 50% of all ALS patients exhibit cognitive impairment, with around 15% developing overt FTD or exhibiting co-morbidity at onset^4,5^. Similarly, approximately 40% of all FTD patients show motor symptoms, with 12% meeting the criteria for ALS^6,7^. While pathogenic variants in *SOD1* and *MAPT, GRN* cause ‘pure’ ALS^8^ or ‘pure’ FTD^9^, respectively, pathogenic variants in *C9orf72*, *TARDBP, OPTN, TBK1,* and *UBQLN2* may cause ALS or FTD or comorbid ALS/FTD^10^, making it unclear to what extent phenotypic heterogeneity is determined by the specific causative variant within those genes.

In addition to genetic predisposition, an emerging contributor to the pathogenesis of ALS/FTD is biological sex and, more specifically, sex chromosome genotype. ALS disproportionately affects males, who show higher incidence, prevalence, and mortality of ALS from all causes in almost all studies, with male-to-female prevalence ratios between 1.2-3.0:1^5,11–14^. Clinically, spinal (limb) and respiratory onset is more frequent in males, with bulbar onset (speech and swallowing) predominating in females of older age^15,16^. Further, most studies report a younger age of disease onset in males than females, with the greatest differences in patients 45 and younger^14,15^. This suggests that there are sex-dependent modifiers of ALS risk and progression, which might include environmental factors, sex hormones, or genetic modifiers on sex chromosomes. Indeed, striking differences in phenotype are seen between males and females with *UBQLN2-*linked ALS/FTD, in which the primary causative gene is on the X chromosome^17–22^.

Across several family and cohort studies, patients with *UBQLN2*-linked ALS/FTD show earlier disease onset in males^17,19,20,22^. *UBQLN2*, being subject to X-inactivation, is expressed differently according to sex^23–26;^ in males, the maternally-derived and only copy of the X-chromosome is expressed in all cells, while females have a mosaic of cells expressing either the maternal or paternal X allele^27,28^. Such genetic diversity in females is proposed to offer resilience against a range of X-linked neurological diseases^29^, potentially delaying or preventing onset altogether^30–32^. This suggests that female resilience to ALS/FTD may impact pathology, phenotype, and clinical features other than onset, and that pronounced female resilience might implicate a *UBQLN2* variant rather than a non-X-linked determinant in causing disease^33,34^.

*UBQLN2-*linked ALS/FTD is rare, affecting between 1-4% of all ALS cases, with most reports being small cohort or case studies^35^. Consequently, determining the extent to which specific *UBQLN2* variants and patient sex influence disease onset, pathology, phenotype, and other clinical features requires meta-analysis. Here we analysed published reports of individuals with *UBQLN2* variants who presented with ALS, FTD, other forms of motor neuron disease (progressive bulbar palsy (PBP) or primary lateral sclerosis (PLS)), or hereditary spastic paraplegia (HSP), as well as gene-positive asymptomatic relatives, to investigate the relationships between disease features, *UBQLN2* variants, and patient sex. This included examination of ubiquilin 2 neuropathology in brain tissue according to *UBQLN2* variant and patient sex. We find that ALS/FTD manifests in several ways and differs between *UBQLN2* variants depending on variant pathogenicity.

## Materials and methods

### Meta-analysis of ALS cases harbouring *UBQLN2* variants

Journal articles were identified in PubMed (January 1993-July 2024) using the search terms *UBQLN2*, ubiquilin 2, motor neuron disease, amyotrophic lateral sclerosis, and ALS, combined with clinical, variant(s), mutation(s), neuropathology, tissue, or immunohistochemistry. Articles lacking clinical information, or that were not primary research articles, were excluded. Individuals were included who i) harboured *UBQLN2* genetic variants and had ALS/FTD, ii) were inferred to harbour *UBQLN2* variants as ALS/FTD-affected members of *UBQLN2*-linked ALS/FTD families or iii) harboured *UBQLN2* variants as members of *UBQLN2*-linked ALS/FTD families but were asymptomatic at the time of publication. At the time of publication, several gene-positive family members of the identified affected individuals were reported to have other forms of MND, namely PBP) or PLS or HSP, a common ALS-mimic. Because *UBQLN2*-linked ALS/FTD exists on a clinical spectrum and these other MNDs can evolve into ALS as their disease progresses, these individuals were also included. Data from 27 articles were extracted, and updated information was requested from the corresponding authors.

Clinical data, including the age of disease onset, disease duration, age at death, site of onset, and acquired symptoms, were extracted. Disease onset was classified by site (spinal, spinal – upper limb, spinal – lower limb, bulbar, cognitive). Clinical symptoms throughout disease were tabulated and categorised into motor, cognitive, and behavioural and psychological symptoms. Where possible, symptoms were separated as to whether spinal-onset ALS affected the upper motor neurons (UMNs, manifesting as weakness, hyperreflexia, and spasticity) or lower motor neurons (LMNs, weakness, atrophy, and fasciculations). Bulbar symptoms were defined as those affecting speech (dysphagia and aphasia) or swallowing (dysarthria). Cognitive features, including impairments in memory, executive function, and intellect, as well as behavioural and psychological impairments including disinhibition, apathy, and compulsivity, were recorded. Biological sex was also documented. Duplicate data was observed for two studies (variant p.Ser340Ile, family UBQLN2#4^36,37^) and was combined in analysis.

### Bioinformatic variant analysis

The annotated human (hg38) reference gene for *UBQLN2* [NM_013444] was used to retrieve PhyloP evolutionary conservation scores through alignment with 14 species for each variant position from the UCSC Genome Browser. The sequence of ubiquilin 2 protein [NP_038472] was retrieved from the NCBI Protein database. To assess the evolutionary conservation of encoded amino acids, peptide sequences orthologous to the human ubiquilin 2 protein were obtained from 11 mammalian species, representing each of the major evolutionary branches, and these were aligned using ClustalW2 (v1.2.4).

Variant minor allele frequencies were obtained from gnomAD v3.1.2 (non-neuro cohort). The following tools were used to predict variant pathogenicity *in silico*; Functional Analysis through Hidden Markov Models v2.3 (FATHMM)^38^, Combined Annotation-Dependent Depletion (CADD) v1.7^39^, MutationTaster2021^40^, PolyPhen2 v2.2.8^41^, MutPred2.0^42^, SIFT 4G^43^, PMUT^44^ and REVEL^45^. Predictions were collated per variant and tabulated in Supplementary Table 3. Pathogenicity classifications (‘pathogenic’, ‘likely pathogenic’, ‘benign’, ‘likely benign’, or of ‘uncertain significance’) of single and small nucleotide *UBQLN2* variants identified in the meta-analysis were retrieved from NCBI dbSNP, ClinVar, and UniProt databases, each of which classifies variants according to American College of Medical Genetics and Genomics and the Association of Molecular Pathology (ACMG/AMP) guidelines^46^. Benign variants were defined as either “likely benign” or “benign” variants and similarly, pathogenic variants were defined as either “likely pathogenic” or “pathogenic”. Using these databases, many variants had conflicting assessments (18.6%; 8/43) or no assessment (46.5%; 20/43). Therefore, all variants underwent appraisal per ACMG/AMP guidelines using InterVar^47^, drawing upon minor allele frequency, presence in controls, *in vitro* or *in vivo* functional studies showing a deleterious or neutral effect, and segregation within >1 affected family members for a combined pathogenicity classification used for further analysis (Supplementary Table 3).

### Meta-analysis statistical analysis

Outlier detection within age at onset, age at death, and disease duration data was performed using a Grubbs’ analysis with Alpha = 0.0001, using GraphPad Prism. One outlier was identified in the disease duration data using Grubbs’ method (G = 7.422; individual II:3 from Fam C harbouring a p.Pro506Ala variant) and excluded from analysis. Sex-specific age at onset, disease duration, and age at death differences were assessed using a two-tailed Mann-Whitney test. A two-way ANOVA with Šídák’s multiple corrections test assessed sex-specific differences in family-matched data. Sex-specific differences in age at onset, according to the ubiquilin 2 domain, were computed using multiple *t*-tests corrected for comparisons using the Holm-Šídák method. Following Kaplan-Meier survival curve plotting, log-rank tests were used to estimate sex differences in the median disease duration. Per sex, chi-squared tests were used to compare the site of onset distributions.

### CNS tissue and fluorescent immunohistochemistry

Formalin-fixed paraffin-embedded (FFPE) post-mortem motor cortex, spinal cord, and hippocampal tissue from a female *UBQLN2-*linked (p.Thr487Ile) ALS/FTD case (MN17) was collected and processed in the New Zealand Human Brain Bank (NZ HuBB) following guidelines previously described^48^ but with fixation for 7 years. Long- (9.25 years) and normally fixed (7 days) tissues were also obtained from a single sporadic female ALS/FTD case (MN15; age at onset = 52) from the NZ HuBB. Despite prolonged fixation of MN17 and MN15, the antigenicity of TDP-43 aggregates was unaffected compared to normally fixed MN15 tissue (Supplementary Figure 1). Unfortunately, no paired long- and normally fixed tissue was available for comparison within any case that had ubiquilin 2 aggregates. Additional FFPE hippocampal tissue from two other affected family members of MN17 (both carrying the *UBQLN2* p.Thr487Ile variant, were obtained from Macquarie University Motor Neuron Disease Research Centre (pedigree ID IV:10 in Figure 1A of ^49^, with ALS) and the Victoria Brain Bank (pedigree ID V:7 in Supplementary Figure 1 of ^3^, with ALS-FTD). FFPE hippocampal tissue from a progressive supranuclear palsy (PSP) case carrying a *UBQLN2* p.Ser222Gly variant of uncertain significance was obtained from the London Neurodegenerative Diseases Brain Bank and Brains for Dementia^50^. All clinical and neuropathological diagnoses were conducted as described previously^51,52^.

**Figure 1.**
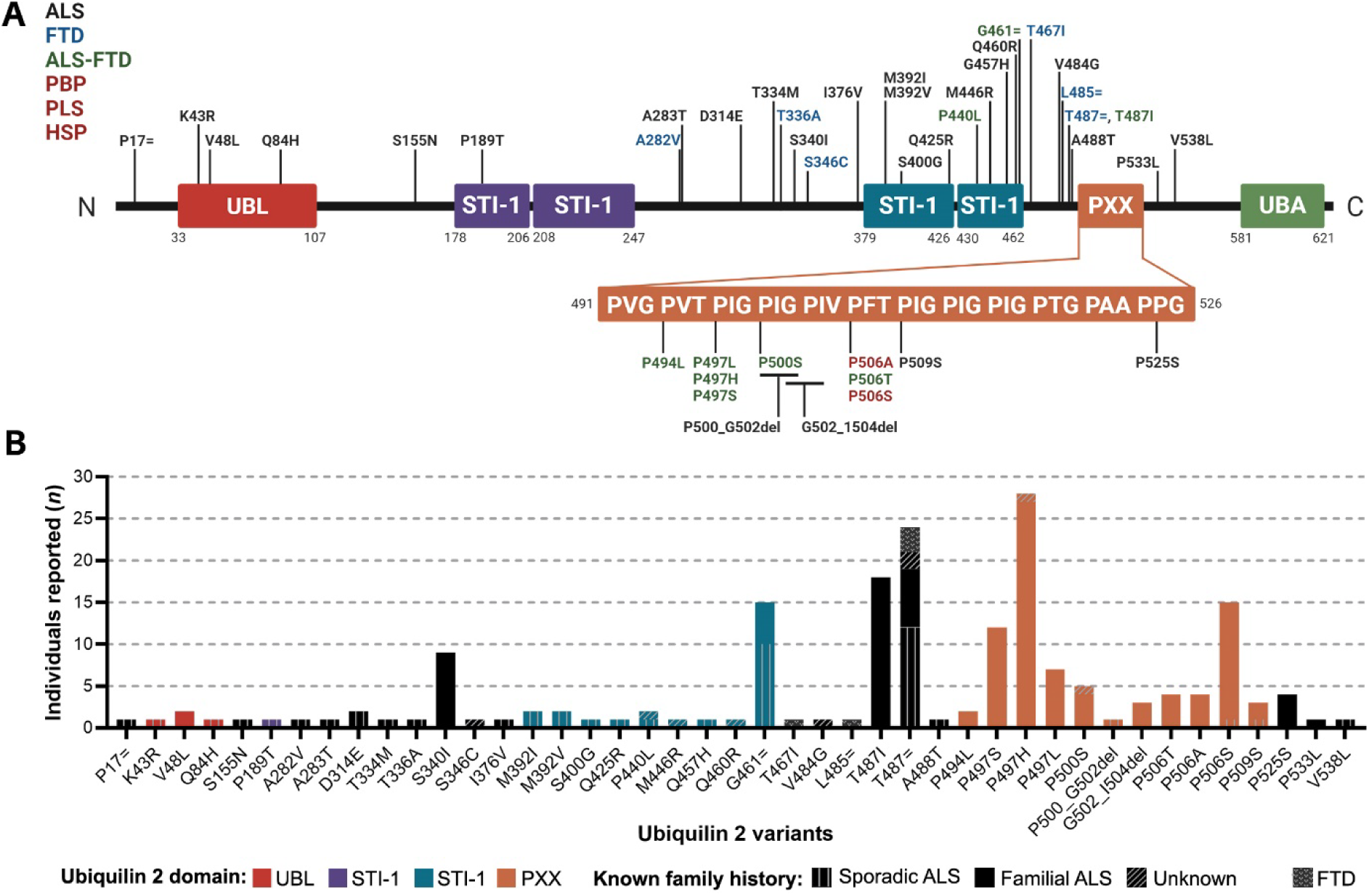
Locations and frequencies of ubiquilin 2 variants in ALS/FTD individuals and families. (**A**) Predicted structural and functional domains of ubiquilin 2 showing the 43 unique variants identified in individuals affected with ALS (black), FTD (blue), ALS-FTD (green), or with ALS or FTD who also had family members who manifested with PLS, PBP, or HSP, all shown in red. (**B**) Variants clustered in the PXX domain (orange bars) are reported in the greatest number of individuals. Box colours (**A**) and bar colours (**B**) correspond to ubiquilin 2 protein functional domains, as per labels in (**A**). Variants are further classified into whether affected individuals had known family history (fALS, filled boxes) or not (sALS, vertical lines) or whether this was not reported or investigated (unknown, diagonal lines). Variants found exclusively in individuals with FTD are denoted by the filled check pattern. Abbreviations: ALS, amyotrophic lateral sclerosis; FTD, frontotemporal dementia; HSP, hereditary spastic paraplegia; PBP, progressive bulbar palsy; PLS, primary lateral sclerosis; PXX, proline-rich domain; STI-1, stress-induced 1 domains; UBA, ubiquitin-associated domain; UBL, ubiquitin- like domain.

Fluorescent immunohistochemistry was performed as previously described^52,53^. Briefly, primary antibody mouse IgG2α anti-ubiquilin 2 (Santa Cruz Biotechnology, sc100612, RRID: AB_2272422) was diluted 1:800 in 1% normal goat serum and applied to sections overnight before washing. Secondary antibody goat anti-mouse IgG 594 (Jackson ImmunoResearch, 115- 585-166, RRID: AB_2338883) was diluted 1:500 in 1% normal goat serum and applied to sections. Nuclei for MN17 and IV:10 were visualised with Hoechst 33342 (1:2000, H3750) stain before washing. V:7 showed poor Hoechst nuclei labelling, therefore a primary antibody against histone H3 (Abcam, ab1791, 1:500, RRID: AB_302613) was used as a nuclear marker prior to secondary (Sigma-Aldrich, goat anti-rabbit IgG biotin, B7389, 1:500, RRID: AB_258613) and tertiary (ThermoFisher Scientific, Streptavidin 405, S32351, 1:200) labelling. Images of coverslipped sections were acquired using a Zeiss Z2 Axioimager with a MetaSystems VSlide slide scanning microscope (20x, 0.9 NA) with a Colibri 7 light source. Images were extracted using VSViewer software (MetaSystems, v.1.1.106) and processed in Adobe Photoshop (version 25.5.1).

### Quantitative analysis of ubiquilin 2 aggregates

Image analysis methods to quantify relative ubiquilin 2 aggregate deposition were developed in MetaMorph v7.10.5.476 (Molecular Devices), as previously described^52^. Sub-regions of interest were identified using nuclei staining. Ubiquilin 2 aggregates were segmented using a consistent absolute threshold labelling intensity within each case, excluding single positive pixels, to create a mask isolating single aggregates.

## Results

### *UBQLN2* variants in ALS/FTD and their predicted pathogenicities

Meta-analysis identified 43 unique *UBQLN2* coding variants in 186 symptomatic individuals with ALS/FTD themselves or in their family and in 51 asymptomatic gene-positive individuals (Table 1, Supplementary Tables 1 and 2). Of these, 36.6% were male, 45.7% were female, and 17.7% did not have biological sex reported. Most individuals were part of cohort and case studies conducted in European (non-Finnish) populations (51.66%), with others conducted in North American (29.2%), East Asian (9.3%), and Middle Eastern (9.9%) populations (Table 1).

**Table 1.**
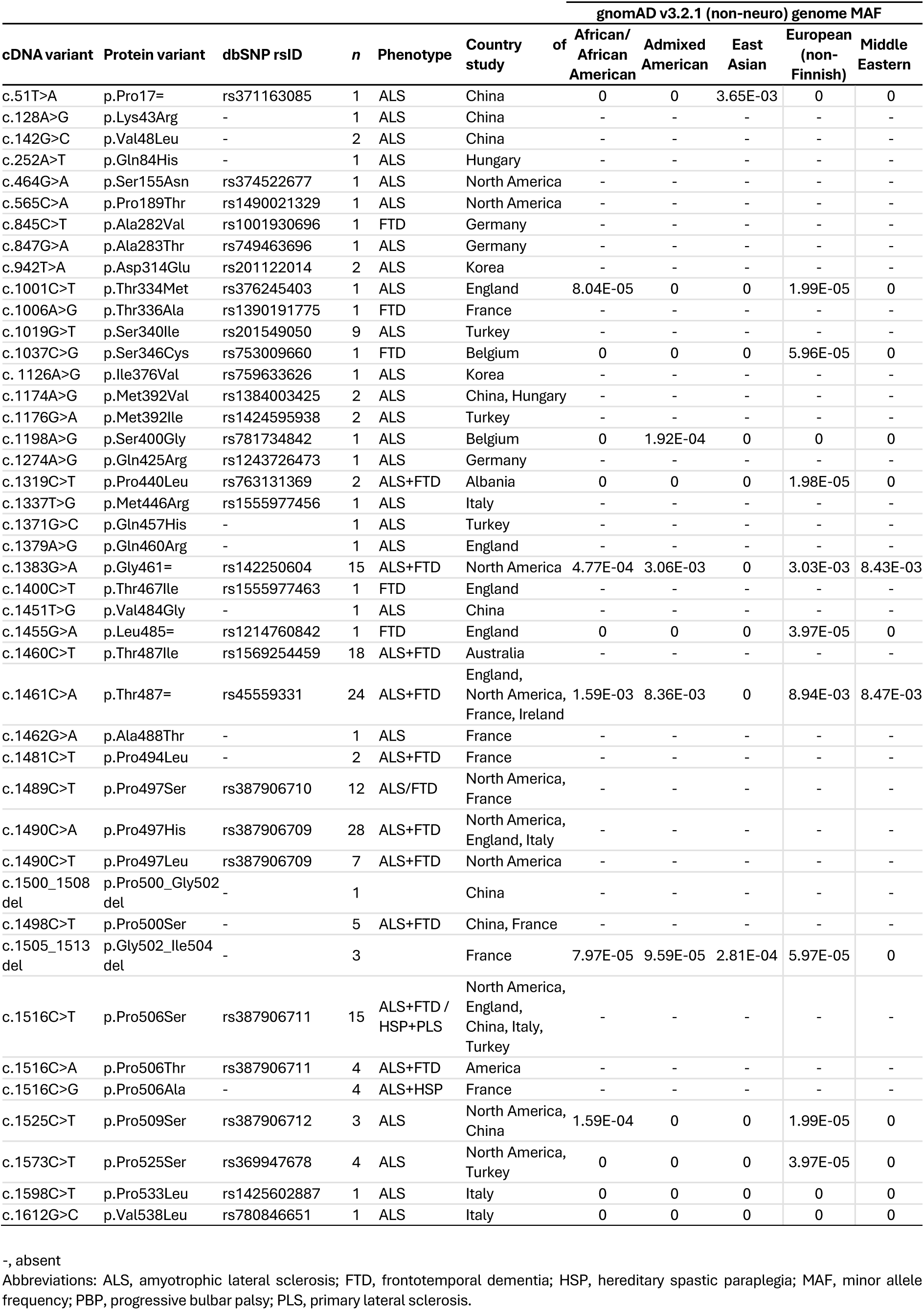
*UBQLN2* [NM_013444] gene variants reported in ALS/FTD and the relevant population frequencies.

Most *UBQLN2* variants were missense (86.0%), with others being synonymous (9.3%) or deletion variants (4.7%, Table 1). These variants primarily affected PXX domain residues, particularly proline 497 (Figure 1A and B). 16/43 variants were reported in >1 familial ALS/FTD case, 27/43 *UBQLN2* variants were found exclusively in sporadic cases, or cases with unknown inheritance, and 4/43 variants were found in cases with ‘pure’ FTD. The most common variant was p.Pro497His segregating in 28 individuals from 3 families^17,19,54–57^ and absent from gnomAD. The p.Thr487Ile variant was also notable, being reported in 18 related individuals across two related families^3,49,52^.

Sequence alignment of *UBQLN2* showed high conservation among placental mammals (Figure 2A), particularly in protein domains affected by disease-causing variants, with 86% of altered amino acids being conserved (Figure 2B). Conservation of amino acids affected by genetic variants was comparable between sporadic and familial cases with 92.0% and 80.0%, respectively.

**Figure 2.**
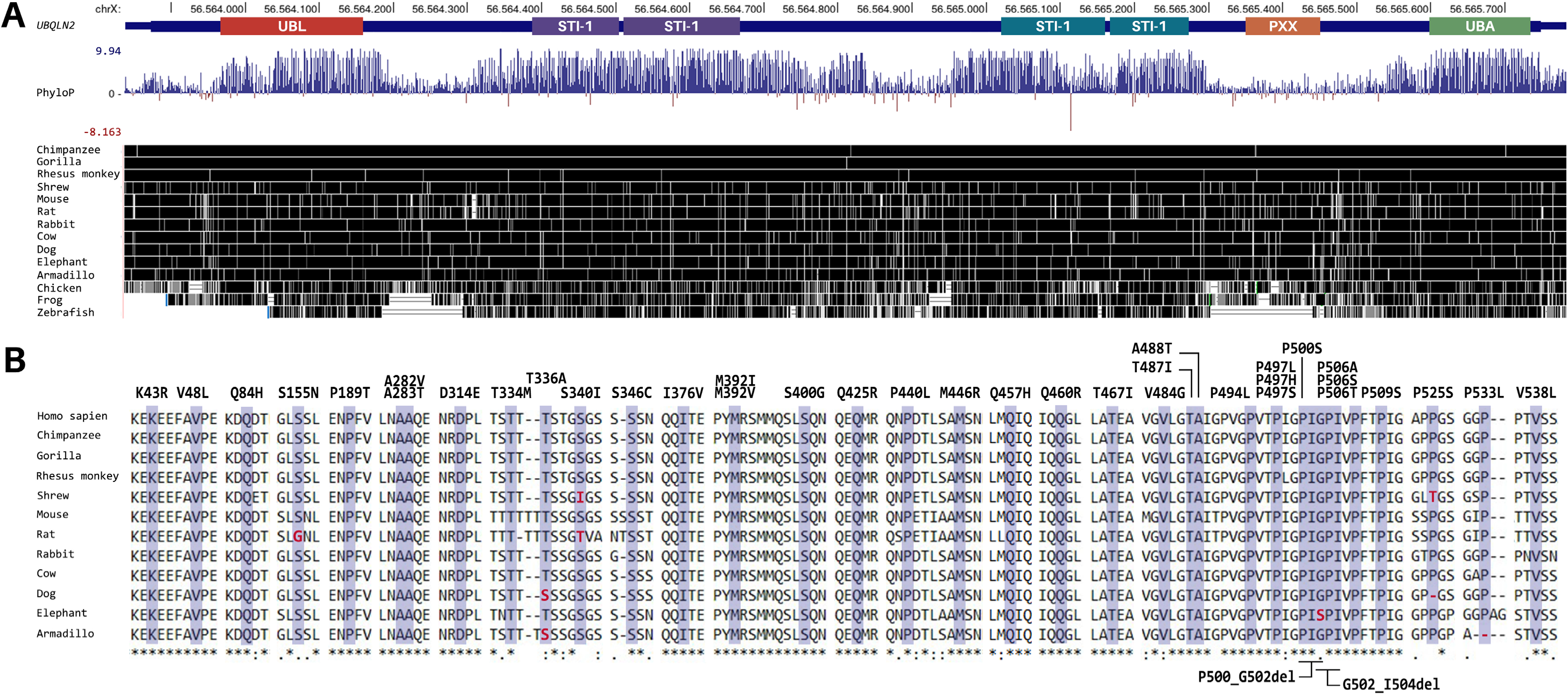
***UBQLN2* variants cause amino acid substitutions and deletions of highly conserved residues.** (**A**) Basewise conservation phyloP scores show a high level of conservation between *UBQLN2* in humans and 14 different mammalian, bird, amphibian, and fish species. Positive scores [blue] indicate conserved sites while negative scores [red] indicate sites predicted to be fast evolving. (**B**) Identified *UBQLN2* variants cause amino acid substitutions in highly conserved protein regions of 11 placental mammalian species and human ubiquilin 2. Sequences used included human (NP_038472.2), chimpanzee (XP_001148687.1), gorilla (XP_004064307.1), rhesus monkey (XP_014983041.1), shrew (XP_006162675.1), mouse (NP_061268.2), rat (NP_001101721.1), rabbit (XP_002720081.1), cow (NP_001192611.1), dog (XP_038306060.1), elephant (XP_010598224.1), and armadillo (XP_012377074.1). Ubiquilin 2 amino acid variants are highlighted in blue. Synonymous variants p.Pro17=, p.Gly461=, p.Leu485=, and p.Thr487= are not shown. Non-conserved variant residues are shown in red. Alignment consensus symbols indicate the degree of conservation: (*), residues identical in all sequences; (:), conserved substitutions; (.), semiconserved substitutions.

### Genetic and clinical analysis of identified *UBQLN2* variants

For symptomatic individuals harbouring any *UBQLN2* variant, the mean age of disease onset was 43.68 ± 17.29 y (range 4-78 y), mean disease duration was 53.89 ± 45.22 months (range 6-204 months), and mean age at death was 50.00 ± 15.25 y (range 18-76 y) (Supplementary Table 1). Spinal onset, including upper limb and lower limb symptoms, was observed in 38/83 individuals (45.8%; 23 male, 15 female). Bulbar onset was seen in 32/84 individuals (38.1%), of whom 12 were male, 19 were female, and one individual whose biological sex was not specified. Cognitive onset affected 14 individuals (4 male, 10 female), 16.7% of the total cohort (Supplementary Table 1).

Clinical phenotypes of symptomatic individuals harbouring any *UBQLN2* variant were variable (Supplementary Table 4). Among 186 individuals, 147 (79.0%) had ALS, 25 (13.4%) had ALS-FTD, and 8 (4.3%) had FTD^21,55,58,59^. The remaining 6 (3.2%) presented with HSP, HSP/ALS, PLS, or PBP. Notably, 63.6% of individuals with FTD (co-morbid or pure) had variants affecting the PXX domain, particularly p.Pro497.

Significant inter-individual variability was observed in clinical progression. For example, *UBQLN2* p.Ser340Ile carriers manifested with spinal motor and intellectual features in 2 related individuals (Supplementary Table 4, UBQLN2#3, individuals II:2 and V:2) but with bulbar onset with progressive cognitive impairment in an unrelated individual (Supplementary Table 4, UBQLN2#4, individual III:12). Variant pathogenicity did not consistently predict site of onset or progression of disease, as seen with variants p.Thr487Ile within families FALS5 and FALS14 and p.Pro497Leu within family 1T or across families e.g. p.Pro497His in individual AB489 versus in family UBQLN2 1 (Supplementary Table 2). Variants affecting p.Pro506 showed particularly diverse phenotypes, including cases of pure HSP, ALS-HSP, PLS, and PBP, with family members often having ALS or ALS+FTD^17,20,22,54^ (Supplementary Table 1).

### Onset of ALS/FTD was earlier in male *UBQLN2* variant carriers than in females

For symptomatic individuals harbouring any *UBQLN2* variant, males showed an earlier mean age at onset than females of 14.89 years (p<0.0001; Figure 3A). This sex disparity was driven by pathogenic (LP/P) *UBQLN2* variants, where males had a mean age at onset 18.15 years earlier than females (p<0.0001; Figure 3B), while no significant differences were observed between the sexes for individuals carrying variants of uncertain significance (VUS) or likely benign/benign (LB/B) variants (Figure 3C and D). The age at onset of both males and females harbouring VUS variants was bimodal, suggesting a mix of true variant pathogenicities. When stratifying pathogenic (LP/P) *UBQLN2* variants by ubiquilin 2 protein domains, this sex-specific difference in age at onset was observed in domains where sex-matched data was available but only statistically significant in the PXX domain, with males developing symptoms on average 18.70 years earlier (p<0.000001; Figure 3E). In contrast, variants of uncertain significance (VUS) stratified by ubiquilin 2 protein domain showed no statistically significant sex-specific differences in age at onset in any *UBQLN2* functional domain, even the ‘hotspot’ PXX domain (p=0.983619, Figure 3F). Within families harbouring pathogenic variants, all variants (11/11) showed earlier onset in males by an average of 18.05 years. However, this difference was statistically significant only in a family with a p.Pro497Leu variant (p=0.0131, n=3 males, 4 females), which likely reflects underpowering for other variants (Figure 3G, Supplementary Table 5). In contrast, more heterogeneity was observed between 3 families harbouring variants of uncertain significance; a related male and female carrying a p.Pro525Ser variant showed onset within a year of each other at 70 and 71, respectively, while the remaining two families showed earlier age at onset in females. However, no relationship was statistically significant (Figure 3H).

**Figure 3.**
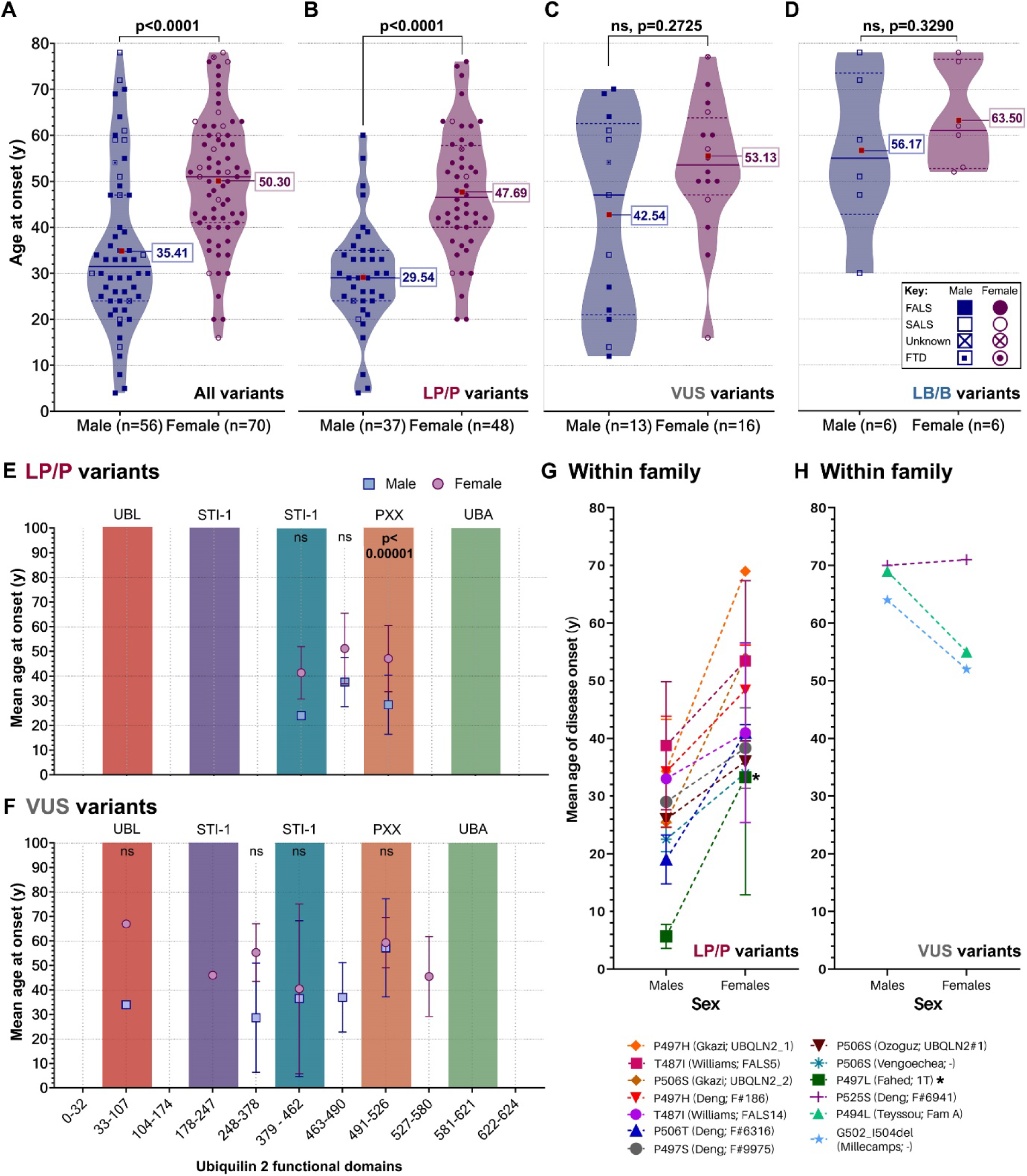
Age of ALS and/or FTD onset is significantly later in females than males carrying pathogenic *UBQLN2* variants. (**A**) Violin plots showing age of disease onset in males and females harbouring any *UBQLN2* variant (All variants), (**B**) likely pathogenic and pathogenic (LP/P) variants, (**C**) variants of uncertain significance (VUS), and (**D**) likely benign or benign (LB/B) variants. Mean indicated by labelled red square. Solid line on violin plot shows median and dashed lines show quartiles. Data points represent individuals and key within figure denotes known ALS/FTD family history. Age at onset was compared using Mann–Whitney tests. (**E**) Age at onset was compared between males and females across ubiquilin 2 domains (colours correspond with protein domains in Figure 1) in individuals harbouring LP/P *UBQLN2* variants, and (**F**) VUS. Values indicate mean ± SD. Age at onset was compared using multiple *t*-tests corrected for multiple comparisons by Holm-Šídák method. (**G** and **H**) Per variant, ages at onset from 13 families with affected male and female members harbouring (**G**) LP/P variants or (**H**) VUS variants were compared using an ordinary two-way ANOVA with Šídák’s multiple comparisons test. Variants are shown in legend below graph with first author of associated publication. Values indicate mean ± SD. Abbreviations: FALS, familial ALS; FTD, frontotemporal dementia; ns, not significant; SALS, sporadic ALS.

### Duration of disease does not differ between affected males and females

For individuals harbouring pathogenic (LP/P) *UBQLN2* variants, median survival time was no different between males (43.00 months [95% CI 43.0-85.9]) and females (48.00 months [95% CI 41.7-68.1]) (p=0.6400; Figure 4A). Earlier mean age at death in males corroborated their earlier mean age at onset, with males dying at a mean age of 39.10 years compared to 51.08 years for females (p<0.0001, Figure 4B). As shown in Figure 4C, spinal onset was the most common site of onset in males harbouring pathogenic *UBQLN2* variants, found in 12/22 males (54.5%), while females tended to present with bulbar symptoms initially (13/25 females, 52.0%). Cognitive onset was rare, being seen in only 1/22 males (4.5%) and 4/25 females (16.0%).

**Figure 4.**
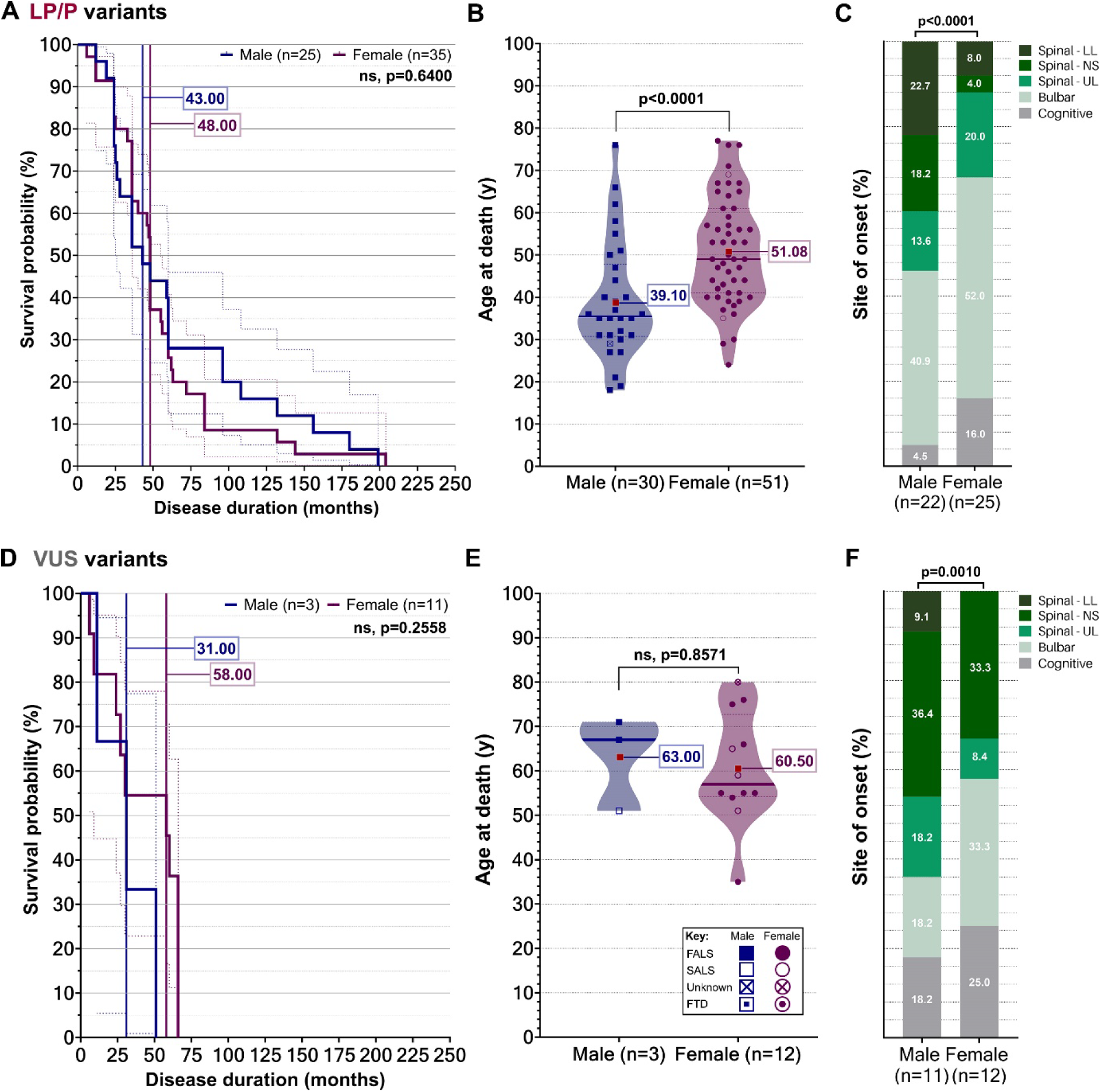
Disease duration does not differ between males and females carrying any *UBQLN2* variant or a pathogenic variant. In affected individuals harbouring likely pathogenic or pathogenic (LP/P) *UBQLN2* variants (**A**) disease duration, (**B**) age at death, and (**C**) site of onset were compared between males and females. In affected individuals harbouring *UBQLN2* variants of uncertain significance (VUS), (**D**) disease duration, (**E**) age at death, and (**F**) site of onset were compared. Differences in survival were assessed using log-rank Mantel-Cox tests. Vertical lines show median survival time per sex. Data points represent individuals and key within figure denotes known ALS/FTD family history. Age at onset was compared using Mann–Whitney tests. Differences in age at death were assessed using a Mann-Whitney test. On violin plots, mean indicated by labelled red square. Solid line shows median and dashed lines show quartiles. Abbreviations: FALS, familial ALS; FTD, frontotemporal dementia; ns, not significant; spinal – LL, spinal – lower limb; spinal – NS, spinal – not specified; spinal – UL, spinal – upper limb; SALS, sporadic ALS.

For individuals with variants of uncertain significance, survival times were not different between sexes (males: 31.00 months [95% CI -9.6-71.6]) females: 58.00 months [95% CI 27.8- 59.1], p=0.2558, Figure 4D). Further, age at death for these individuals was not significantly different between males (63.00 years) and females (60.50 years) (p=0.8571; Figure 4E). Site of onset patterns were consistent with that of pathogenic variants, with males typically showing spinal onset and females showing bulbar onset (Figure 4F).

No significant sex differences were observed in age at onset per ubiquilin 2 protein domain, site of onset, survival, or age at death for likely benign/benign variants (Supplementary Figure 2). All sex-specific differences are summarised in Table 2.

**Table 2.**
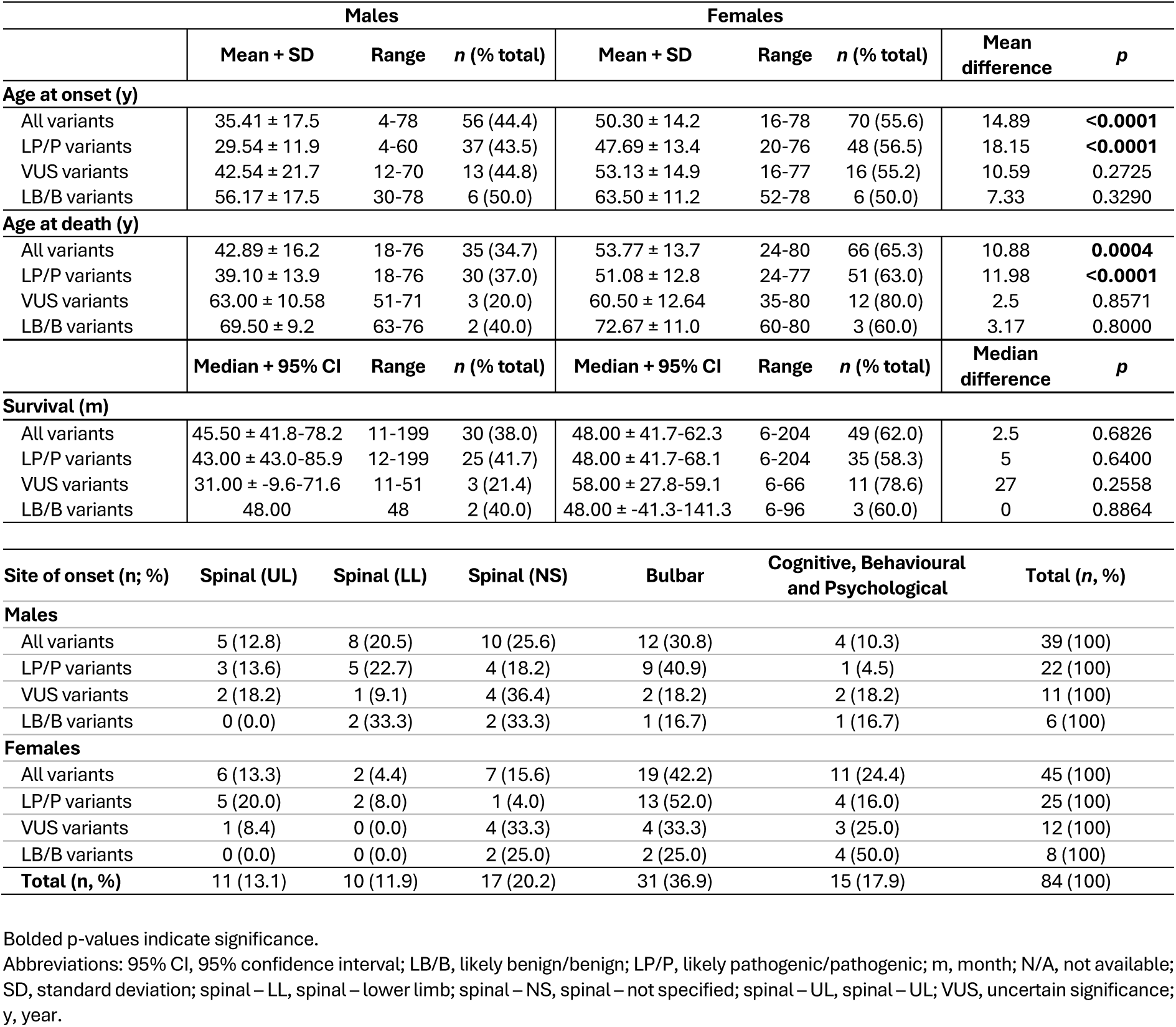
Sex-specific clinical differences in *UBQLN2*-linked ALS/FTD.

**Table 3.** Demographic and clinical information of 3 related *UBQLN2* p.Thr487Ile cases and an unrelated *UBQLN2* p.S222G case with immunohistochemistry data.

| Case ID | Genetic mutation | Diagnosis | Sex | AAO (y) | DD (m) | AAD (y) | PMD (h) | SOO |
| --- | --- | --- | --- | --- | --- | --- | --- | --- |
| MN17 – T487I | *UBQLN2* c.1460C>T (p.T487I) | MND-ALS+FTD | F | 58 | 36 | 61 | 69 | Spinal |
| IV:10 – T487I | *UBQLN2* c.1460C>T (p.T487I) | MND-ALS | M | 34 | 12 | 35 | - | Bulbar |
| V:7 – T487I | *UBQLN2* c.1460C>T (p.T487I) | MND-ALS+FTD | M | 30 | 108 | 39 | 25.5 | - |
| BBN_16359 | *UBQLN2* c.664A>G (p.S222G) | PSP | M | 93 | - | - | 19.5 | - |

Abbreviations: AAO, age at onset; DD, disease duration; AAD, age at death; h, hour; m, month; PMD, post-mortem delay; PSP, progressive supranuclear palsy; SOO, site of onset; y, year; -, unknown.

### Males with a pathogenic *UBQLN2* p.Thr487Ile variant showed increased ubiquilin 2 protein neuropathology compared to a related carrier female

Fluorescent labelling in the hippocampus of three related p.Thr487Ile cases (2 male, 1 female) revealed that cases V:7–T487I (M) and IV:10–T487I (M) had many more ubiquilin 2 aggregates than case MN17–T487I (F) in the motor cortex, spinal cord, and hippocampus (Figure 5). Although fixation of the female tissue had been greatly extended, the antigenicity of protein aggregates was preserved in a comparator ALS/ FTD case for which both long- and short-fixed tissue was available (Supplementary Figure 1).

**Figure 5.**
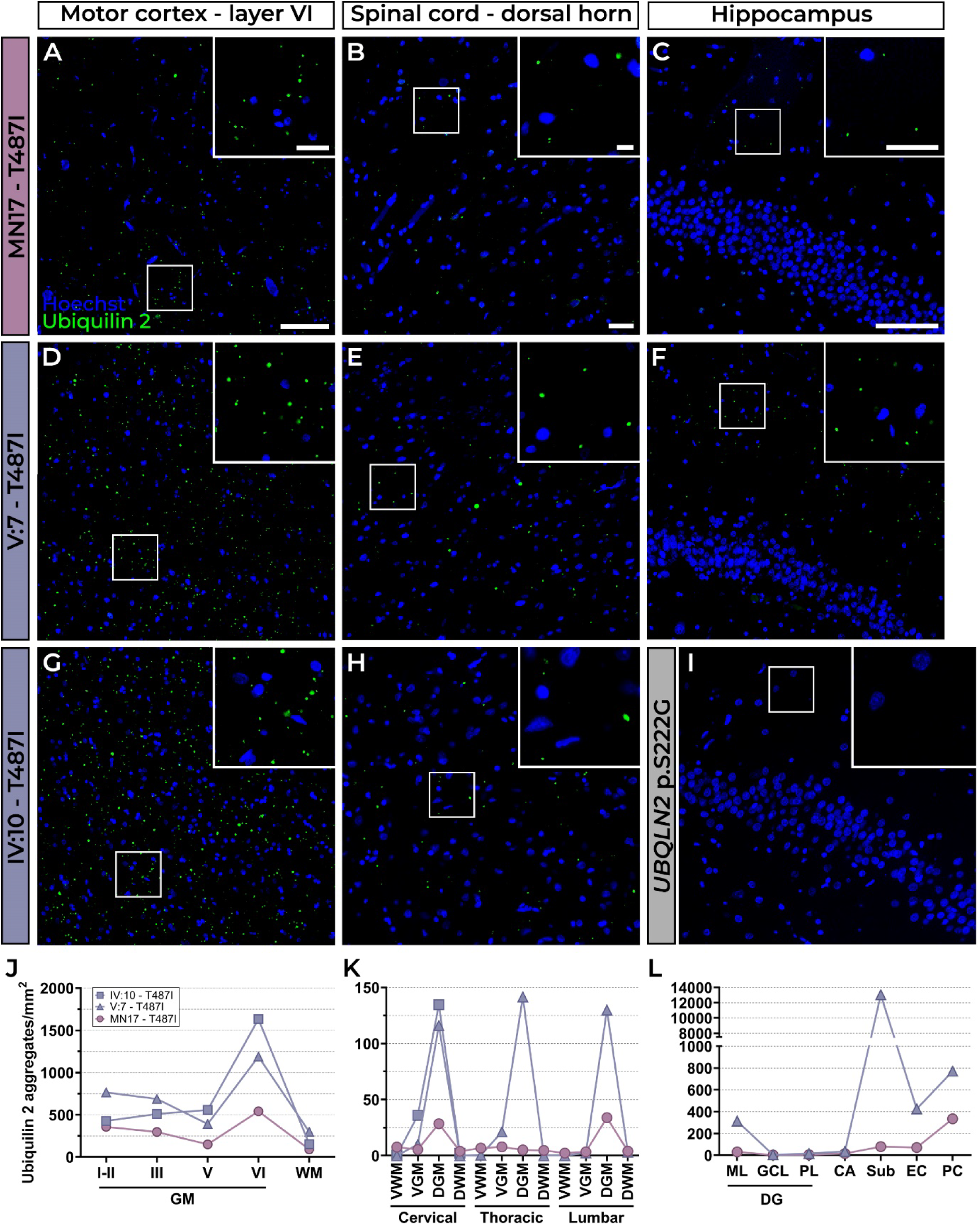
Differential ubiquilin 2 aggregate load in male and female *UBQLN2* p.T487I- linked ALS/FTD. Load of ubiquilin 2 aggregates, shown in green, was compared between three related individuals with ALS-FTD harbouring a pathogenic *UBQLN2* p.Thr487Ile mutation, in the motor cortex (**A, D, G**, quantified in **J**), spinal cord dorsal horn (**B, E, H**, quantified in **K**), and hippocampus (**C, F**, quantified in **L**). Note: hippocampal tissue was not available for IV:10 – T487I. (**I**) A case harbouring a *UBQLN2* p.Ser222Gly variant of uncertain significance and presenting with progressive supranuclear palsy showed no ubiquilin 2 pathology. Scale bar in main images is 100 µm, and 25 µm in zooms.

In the motor cortex layer VI, ubiquilin 2 aggregate densities were 1188.94 aggregates/mm^2^ for male relative V:7-T487I, and 1634.38 aggregates/mm^2^ for IV:10-T487I, compared to 541.45 aggregates/mm^2^ for MN17-T487I (female) (Figure A, D, G, and I). In the cervical dorsal grey matter, ubiquilin 2 aggregate densities were 116.0 and 134.7 aggregates/mm^2^ for male relatives V:7-T487I and IV:10-T487I, respectively while the female MN17-T487I had 28.54 aggregates/mm^2^ ((Figure 5B, E, H, and J). In the hippocampus, V:7-T487I had a significantly higher aggregate density in the subiculum (13,040.9 aggregates/mm^2^) than MN17-T487I (79.39 aggregates/mm^2^; Figure 5C, F, and K). A PSP case harbouring a *UBQLN2* p.Ser222Gly variant of uncertain significance (Figure 5I) showed no ubiquilin 2 in the hippocampus, further demonstrating the value of neuropathology in determining *UBQLN2* variant pathogenicity.

## Discussion

### Consistent features of pathogenic *UBQLN2*-linked ALS/FTD

*UBQLN2*-linked ALS/FTD shows considerable clinical and pathological heterogeneity, highlighting the complexity of the involvement of ubiquilin 2 in disease pathogenesis but also complicating predictions of *UBQLN2* variant pathogenicity. Although numerous animal studies show that *UBQLN2* pathogenic variants are deleterious to neurons^60–63^, there is substantial variation in disease presentation, even among family members sharing an identical pathogenic variant and particularly between the sexes.

#### UBQLN2-linked ALS/FTD shows earlier onset and greater neuropathology in males than females

A key finding is the significantly earlier age at onset in males than females with pathogenic *UBQLN2* variants, based on analysis of 86 individuals. This supports and extends previous studies of 54 and 43 individuals, which did not stratify by variant classification and thus the reported sex differences (13.36 and 13.25 years, respectively) may have been underestimates for cases where *UBQLN2* is the major genetic contributor^17,64^. Indeed, in our analysis, males carrying any *UBQLN2* variant typically developed ALS/FTD 14.9 years earlier than females, but this increased to 18.13 years for those harbouring pathogenic variants. Further, the pronounced sex difference in age at onset for variants in the proline-rich PXX domain implicates this region in disease risk, but variants upstream of the PXX domain also showed sex differences in onset, indicating that the PXX domain is not wholly contributory to the sex-specific differences observed, as previously asserted^17^.

Male susceptibility to early onset of *UBQLN2*-linked ALS/FTD likely derives from hemizygosity of the mutant allele, resulting in expression of mutant ubiquilin 2 in every cell, in contrast to females who are heterozygous for the mutant allele, resulting in mosaic expression of wildtype and mutant ubiquilin 2 by females^32,65^. Indeed, ubiquilin 2 aggregate load in post-mortem human brain tissue was greater in two male cases carrying a pathogenic p.Thr487Ile variant than 1 female case. Although these findings require validation within additional families, they suggest that ubiquilin 2 load is sex-dependent and that ubiquilin 2 misfolding and aggregation may be a driver of disease onset.

The striking sex differences in age at onset were absent from the groups of variants of uncertain significance or likely benign/benign variants, suggesting that most variants in those groups are not pathogenic or their contribution to the genetic architecture of ALS/FTD onset risk is small. Alternatively, these variants may yet be pathogenic, but sex difference analyses were underpowered because they are so rare. Indeed, the age at onset of males harbouring variants of uncertain significance appeared bimodal, so although this variant group is poorly understood through a lack of structural, functional, or pathological examination, it may comprise a mix of true pathogenic variants and true benign variants. Indeed, compared to wildtype, VUS *UBQLN2* p.Pro494Leu lymphoblasts (a variant found in FALS only) impaired autophagy and HSP70 binding^22^. In contrast, turbidity assays of VUS *UBQLN2* p.Val538Leu (variant found in SALS only) showed comparable behaviour to wildtype, providing functional evidence against its pathogenic involvement in ALS/FTD^66^. Conflicting functional studies of p.Pro525Ser showing slightly elevated ubiquitination and neuronal toxicity in a Drosophila model ^67^ but comparable turbidity behaviour to wildtype^66^, and co-segregation in affected FALS individuals^19,37^ suggest it has small pathogenic involvement in ALS/FTD. Further, some individuals in this cohort carried additional variants in other ALS-linked genes (Supplementary Table 1). A lack of reported clinical information from these individuals makes it challenging to determine the independent contributions of each variant to disease or whether they are true oligogenic cases^68^. Functional studies of *UBQLN2* variants of uncertain significance may further aid in their classification, as would the compilation of age at onset data from more individuals, given that sex difference in age at onset is associated with *UBQLN2* variant pathogenicity.

#### Females with UBQLN2 pathogenic variants show incomplete disease penetrance

The presence of many asymptomatic carriers of *UBQLN2* variants (Supplementary Table 2), most of whom are female, suggests incomplete penetrance of *UBQLN2*-linked disease in females. These individuals (1) may not have manifested with disease at the time of report but eventually would, or (2) may harbour non-pathogenic variants and never manifest with disease or (3) be protected by favourable X-chromosome inactivation skew^65,69^. The direction and degree of X-chromosome inactivation skewing has been shown to influence the onset and severity of other X-linked diseases, including Duchenne muscular dystrophy^70^ and Fabry disease^71,72^, and skew may also change with aging^73^. This presents an important caveat to using sex differences to support variant pathogenicity classification; the lack of a sex difference within a family of *UBQLN2* gene carriers could indicate female skew towards expression of the *UBQLN2* variant, rather than lack of pathogenicity of that variant.

#### Females with UBQLN2 pathogenic variants may show bulbar phenotype predisposition

In this cohort, females with *UBQLN2*-linked ALS/FTD were more likely to present with bulbar onset compared to males. Among females harbouring any *UBQLN2* variant, 43.2% had bulbar onset, increasing to 50.0% for those with pathogenic variants. In contrast, 30.8% of males with any variant and 40.9% with pathogenic variants presented with bulbar onset. A study of 600 sporadic ALS patients showed females are more likely to present with bulbar phenotypes than men (37% of females vs 23% of males^74^). However, these numbers are below what is observed in *UBQLN2*-linked ALS/FTD, particularly within males. Speculatively, the selective vulnerability of the medulla in *UBQLN2-*linked ALS/FTD may predispose to bulbar phenotype as ubiquilin 2 aggregates are found in the middle and lower medulla, and in the inferior olivary nucleus^52^. Hexanucleotide repeat expansions in *C9orf72*, a common genetic cause of ALS/FTD, were also found to have a predisposing effect upon bulbar onset through an interaction with increasing age but was not sex-specific^16^. Indeed, as *UBQLN2*-linked ALS/FTD females show later onset, bulbar predisposition may also/instead be a result of increasing age and perhaps pathology propagation within the brain over time. Nevertheless, the compilation of clinical data for more individuals with *UBQLN2-*linked ALS/FTD, as well as further exploration of degeneration across brain regions in these affected individuals, is warranted to clarify relationships between pathology burden, site of onset, increasing age, and sex in *UBQLN2-*linked ALS/FTD.

#### Disease onset and progression may be independent processes

Despite the significant differences in age at onset, disease duration was the same between males and females, irrespective of the classification of their *UBQLN2* variant (Figure 4 and Supplementary Figure 2). Given that *UBQLN2-*linked ALS/FTD age at onset and resulting ubiquilin 2 pathology in tissue is sex-specific but that disease duration is not, the pathomechanisms of disease onset and progression are likely to be distinct. This hypothesis is supported by recent genome-wide association and cohort analysis studies^75,76^, and our work shows that the burden of TDP-43 rather than ubiquilin 2 correlates with neurodegeneration^52^. A similar difference was observed in Huntington’s disease whereby increased CAG repeat length and Htt protein load correlated with earlier age of motor symptom onset but not disease progression rate and survival^77^. The additional determinants or ‘second hits’ that mediate neurodegeneration warrant further exploration, but dysfunction of TDP-43 is likely among them.

#### UBQLN2 variants result in diverse motor and cognitive phenotypes

Even pathogenic *UBQLN2* variants that exhibit complete penetrance may show variable expressivity, as evidenced by cases within the same family where individuals present with different motor neuron diseases or dementia-like disorders. For example, a pathogenic *UBQLN2* p.Pro506Ser carrier had ‘pure’ HSP, while their mother had ALS/FTD, and a sibling had ‘pure’ ALS^17^. Similarly, a *UBQLN2* p.Pro506Ala carrier initially diagnosed with HSP later developed ALS, with relatives showing varied MND presentations^22^. These cases demonstrate the overlapping genetic and mechanistic underpinnings of these MNDs and dementia-like disorders but suggest non-*UBQLN2* determinants of phenotype. Similarly, several *ERLIN2-* linked HSP and *UBQLN2-*linked FTD patients have been reported who did not convert to ALS, indicating that additional modifying factors contribute to specific motor or cognitive phenotypes. Taken together, genetic testing for *UBQLN2* mutations should also be undertaken in individuals with other motor neuron disease phenotypes, such as HSP, as well as dementia-like disorders like FTD to ensure disease is accurately and rapidly diagnosed and moreover, further understand the role of pathogenic *UBQLN2* variants across a broader phenotypic spectrum.

## Conclusion

This is the largest cohort clinical meta-analysis of *UBQLN2-*linked ALS/FTD, with important implications for diagnosis and management. To have confidence that a variant is disease-causing (pathogenic) requires the integration of multiple strands of evidence at the gene-level (conservation impact), protein-level (physicochemical properties, and functional impact), and case-level (inheritance, segregation, predictable or consistent clinical features of disease onset and progression). Despite the heterogeneity of the identified cohort and incomplete data capture, significant sex-specific differences in *UBQLN2-*linked ALS/FTD were found in age at onset, age at death, and pathological burden in brain tissue of related affected individuals. These differences are predictable clinical features of *UBQLN2-*linked ALS/FTD and can be used to support variant pathogenicity classification.

Despite a similar disease duration, males with pathogenic *UBQLN2* variants show earlier disease onset, and increased ubiquilin 2 aggregate burden across the central nervous system than females, likely due to the hemizygous nature of their *UBQLN2* variant. This difference holds across known pathogenic variants and a subset of variants that are currently of unknown significance, which may support their reclassification. Prospective longitudinal studies of such variants are warranted to map disease progression and identify phenotype-specific characteristics of disease, as well as assess these variants in functional studies. Further, this study strengthens the evidence of clinical overlap between ALS and FTD, and rarer motor diseases such as HSP and PSP in the observable disease spectrum for *UBQLN2* pathogenic variants. Ultimately, these results will inform predictions of ALS and ALS/FTD risk in families harbouring a *UBQLN2* variant that may be novel or known. For both patients and clinicians, knowledge of pathogenic sex-specific patterns in clinical presentations of *UBQLN2-*linked ALS/FTD may aid in early-detection, diagnostic accuracy, disease management, and patient outcomes, for those affected and their asymptomatic relatives.

## List of abbreviations

ALS: Amyotrophic lateral sclerosis
FTD: Frontotemporal dementia
HSP: Hereditary spastic paraplegia
MND: Motor neuron disease
PBP: Progressive bulbar palsy
PLS: Primary lateral sclerosis
PSP: Progressive supranuclear palsy
TDP-43: Transactive response DNA binding protein 43 kDa
UBQLN2: Ubiquilin 2

## Acknowledgements

This publication is dedicated to the incredible participants and families who contribute to our research. We thank Marika Eszes at the Centre for Brain Research, University of Auckland, New Zealand and the Neurological Foundation of New Zealand for their ongoing financial support of the Human Brain Bank. We also thank Fairlie Hinton and Dr Catriona McLean at the Victorian Brain Bank, which is supported by The Florey Institute of Neuroscience and Mental Health, The Alfred and the Victorian Forensic Institute of Medicine and funded in part by Parkinson’s Victoria, MND Victoria, FightMND and Yulgilbar Foundation. The imaging data reported in this paper were obtained at the Biomedical Imaging Research Unit (BIRU), operated by the Faculty of Medical and Health Sciences’ Technical Services at the University of Auckland. The authors also wish to thank Drs Stephanie Millecamps, Nailah Siddique, and Kornelia Tripolszki for their personal correspondence and updated information on patient clinical features.

## Acknowledgements and funding

This publication is dedicated to the incredible patients and families who contribute to our research.

## Authors’ contributions

KMT, LRN, MEVS, and KLW conducted or designed experiments and performed data analysis or designed analysis methods; CT conducted neuropathological diagnostics; MD provided study supervision and resourcing; GAN identified the *UBQLN2* patients; RLMF, MAC, and IPB coordinated the banking and use of human tissue for study; KMT, BVD, ELS, and KLW wrote the manuscript; KMT and ELS conceived of and designed the study. All authors read, edited, and approved the final manuscript.

## Data availability

The datasets used and/or analysed in the current study are available within the article and its supplementary material.

## References

1. van Es, M. A. et al. Amyotrophic lateral sclerosis. The Lancet 390, 2084–2098 (2017).

2. Carbayo, Á. et al. Clinicopathological correlates in frontotemporal lobar degeneration: motor neuron disease spectrum. Brain (2024) doi:10.1093/brain/awae011.

3. Thumbadoo, K. M. et al. Hippocampal aggregation signatures of pathogenic UBQLN2 in amyotrophic lateral sclerosis and frontotemporal dementia. Brain 147, 3547–3561 (2024).

4. Burrell, J. R., Kiernan, M. C., Vucic, S. & Hodges, J. R. Motor Neuron dysfunction in frontotemporal dementia. Brain 134, 2582–2594 (2011).

5. Statland, J. M., Barohn, R. J., McVey, A. L., Katz, J. S. & Dimachkie, M. M. Patterns of Weakness, Classification of Motor Neuron Disease, and Clinical Diagnosis of Sporadic Amyotrophic Lateral Sclerosis. Neurol Clin 33, 735–748 (2015).

6. Bersano, E., Manera, U. & Huynh, W. Editorial: Cognitive and Behavioral Features in ALS: Beyond Motor Impairment in ALS-FTD Spectrum Disorders. Front Neurol 13, 1–2 (2022).

7. Abrahams, S. Neuropsychological impairment in amyotrophic lateral sclerosis– frontotemporal spectrum disorder. Nat Rev Neurol 19, 655–667 (2023).

8. Huang, M., Liu, Y. U., Yao, X., Qin, D. & Su, H. Variability in SOD1-associated amyotrophic lateral sclerosis: geographic patterns, clinical heterogeneity, molecular alterations, and therapeutic implications. Translational Neurodegeneration vol. 13 Preprint at 10.1186/s40035-024-00416-x (2024).

9. Seelaar, H. et al. Distinct genetic forms of frontotemporal dementia. Neurology 71, 1220–1226 (2008).

10. Kirola, L., Mukherjee, A. & Mutsuddi, M. Recent Updates on the Genetics of Amyotrophic Lateral Sclerosis and Frontotemporal Dementia. Mol Neurobiol (2022) doi:10.1007/s12035-022-02934-z.

11. Hardiman, O. et al. Amyotrophic lateral sclerosis. Nature Reviews 3, (2017).

12. Petrov, D., Mansfield, C., Moussy, A. & Hermine, O. ALS clinical trials review: 20 years of failure. Are we any closer to registering a new treatment? Front Aging Neurosci 9, 1–11 (2017).

13. Marin, B. et al. Variation in world wide incidence of amyotrophic lateral sclerosis: A meta-analysis. Int J Epidemiol 46, 57–74 (2017).

14. McCombe, P. A. & Henderson, R. D. Effects of gender in amyotrophic lateral sclerosis. Gend Med 7, 557–570 (2010).

15. Blasco, H. et al. Amyotrophic lateral sclerosis: A hormonal condition? Amyotrophic Lateral Sclerosis 13, 585–588 (2012).

16. Chiò, A. et al. ALS phenotype is influenced by age, sex, and genetics: A population-based study. Neurology 94, e802–e810 (2020).

17. Gkazi, S. A. et al. Striking phenotypic variation in a family with the P506S UBQLN2 mutation including amyotrophic lateral sclerosis, spastic paraplegia, and frontotemporal dementia. Neurobiol Aging 73, 229.e5–229.e9 (2019).

18. Fahed, A. C. et al. UBQLN2 mutation causing heterogeneous X-linked dominant neurodegeneration. Ann Neurol 75, 793–798 (2014).

19. Deng, H.-X. X. et al. Mutations in UBQLN2 cause dominant X-linked juvenile and adult-onset ALS and ALS/dementia. Nature 477, 211–215 (2011).

20. Vengoechea, J., David, M. P., Yaghi, S. R., Carpenter, L. & Rudnicki, S. A. Clinical variability and female penetrance in X-linked familial FTD/ALS caused by a P506S mutation in UBQLN2. Amyotroph Lateral Scler Frontotemporal Degener 14, 615–619 (2013).

21. Dillen, L. et al. Explorative genetic study of UBQLN2 and PFN1 in an extended Flanders-Belgian cohort of frontotemporal lobar degeneration patients. Neurobiol Aging 34, 1711.e1–1711.e5 (2013).

22. Teyssou, E. et al. Novel UBQLN2 mutations linked to amyotrophic lateral sclerosis and atypical hereditary spastic paraplegia phenotype through defective HSP70- mediated proteolysis. Neurobiol Aging 58, 239.e11–239.e20 (2017).

23. Lyon, M. F. Sex chromatin and gene action in the mammalian X-chromosome. Am J Hum Genet 14, 135–148 (1962).

24. Lyon, M. F. Gene Action in the X-chromosome (Mus musculus L.). Nature 190, 372–373 (1961).

25. Lyon, M. F. Attempts to test the inactive-X theory of dosage compensation in mammals. Genet Res 4, 93–103 (1963).

26. Disteche, C. M. & Berletch, J. B. X-chromosome inactivation and escape. J Genet 94, 591–599 (2015).

27. Shah, K., McCormack, C. E. & Bradbury, N. A. Do you know the sex of your cells? Am J Physiol Cell Physiol 306, (2014).

28. Wu, H. et al. Cellular Resolution Maps of X Chromosome Inactivation: Implications for Neural Development, Function, and Disease. Neuron 81, 103–119 (2014).

29. Davis, E. J. et al. A second X chromosome contributes to resilience in a mouse model of Alzheimer’s disease. Sci Transl Med 12, 1–36 (2020).

30. Echevarria, L. et al. X-chromosome inactivation in female patients with Fabry disease. Clin Genet 89, 44–54 (2016).

31. Migeon, B. R. X-linked diseases: susceptible females. Genetics in Medicine 22, 1156–1174 (2020).

32. Sun, Z., Fan, J. & Wang, Y. X-Chromosome Inactivation and Related Diseases. Genet Res (Camb) 2022, (2022).

33. Fang, H., Disteche, C. M. & Berletch, J. B. X Inactivation and Escape: Epigenetic and Structural Features. Front Cell Dev Biol 7, 1–12 (2019).

34. Carrel, L. & Willard, H. F. X-inactivation profile reveals extensive variability in X-linked gene expression in females. Nature 434, 400–404 (2005).

35. Renton, A. E., Chiò, A. & Traynor, B. J. State of play in amyotrophic lateral sclerosis genetics. Nat Neurosci 17, 17–23 (2014).

36. Kotan, D., İskender, C., Özoğuz Erimiş, A. & Başak, A. N. A Turkish family with a familial ALS-positive UBQLN2-S340I mutation. Noropsikiyatri Arsivi 53, 283– 285 (2016).

37. Özoğuz, A. et al. The distinct genetic pattern of ALS in Turkey and novel mutations. Neurobiol Aging 36, 1764.e9–1764.e18 (2015).

38. Shihab, H. A. et al. Predicting the Functional, Molecular, and Phenotypic Consequences of Amino Acid Substitutions using Hidden Markov Models. Hum Mutat 34, 57–65 (2013).

39. Kircher, M. et al. A general framework for estimating the relative pathogenicity of human genetic variants. Nat Genet 46, 310–315 (2014).

40. Schwarz, J. M., Cooper, D. N., Schuelke, M. & Seelow, D. Mutationtaster2: Mutation prediction for the deep-sequencing age. Nat Methods 11, 361–362 (2014).

41. Adzhubei, I. A. et al. A method and server for predicting damaging missense mutations. Nat Methods 7, 248–249 (2010).

42. Pejaver, V. et al. Inferring the molecular and phenotypic impact of amino acid variants with MutPred2. Nat Commun 11, (2020).

43. Ng, P. C. & Henikoff, S. SIFT: Predicting amino acid changes that affect protein function. Nucleic Acids Res 31, 3812–3814 (2003).

44. López-Ferrando, V., Gazzo, A., De La Cruz, X., Orozco, M. & Gelpí, J. L. PMut: A web-based tool for the annotation of pathological variants on proteins, 2017 update. Nucleic Acids Res 45, W222–W228 (2017).

45. Ioannidis, N. M. et al. REVEL: An Ensemble Method for Predicting the Pathogenicity of Rare Missense Variants. Am J Hum Genet 99, 877–885 (2016).

46. Richards, S. et al. Standards and guidelines for the interpretation of sequence variants: a joint consensus recommendation of the American College of Medical Genetics and Genomics and the Association for Molecular Pathology. Genetics in Medicine 17, 405–424 (2015).

47. Li, Q. & Wang, K. InterVar: Clinical Interpretation of Genetic Variants by the 2015 ACMG-AMP Guidelines. Am J Hum Genet 100, 267–280 (2017).

48. Waldvogel, H. J. et al. The collection and processing of human brain tissue for research. Cell Tissue Bank 9, 169–179 (2008).

49. Williams, K. L. et al. UBQLN2/ubiquilin 2 mutation and pathology in familial amyotrophic lateral sclerosis. Neurobiol Aging 33, 2527.e3–2527.e10 (2012).

50. Keogh, M. J. et al. Genetic compendium of 1511 human brains available through the UK Medical Research Council Brain Banks Network Resource. Genome Res 27, 165–173 (2017).

51. Scotter, E. L. et al. C9ORF72 and UBQLN2 mutations are causes of amyotrophic lateral sclerosis in New Zealand: a genetic and pathologic study using banked human brain tissue. Neurobiol Aging 49, 214.e1–214.e5 (2017).

52. Nementzik, L. R. et al. Distribution of ubiquilin 2 and TDP-43 aggregates throughout the CNS in UBQLN2 p.T487I-linked amyotrophic lateral sclerosis and frontotemporal dementia. Brain Pathology 1–14 (2023) doi:10.1111/bpa.13230.

53. Waldvogel, H. J., Curtis, M. A., Baer, K., Rees, M. I. & Faull, R. L. M. Immunohistochemical staining of post-mortem adult human brain sections. Nat Protoc 1, 2719–2732 (2006).

54. Gellera, C. et al. Ubiquilin 2 mutations in Italian patients with amyotrophic lateral sclerosis and frontotemporal dementia. J Neurol Neurosurg Psychiatry 84, 183– 187 (2013).

55. Ugwu, F. et al. A UBQLN2 variant of unknown significance in frontotemporal lobar degeneration. Neurobiol Aging 36, 546.e15–546.e16 (2015).

56. McLaughlin, R. L. et al. UBQLN2 mutations are not a frequent cause of amyotrophic lateral sclerosis in Ireland. Neurobiol Aging 35, 267.e9–267.e11 (2014).

57. Morgan, S. et al. A comprehensive analysis of rare genetic variation in amyotrophic lateral sclerosis in the UK. Brain 140, 1611–1618 (2017).

58. Lattante, S. et al. Screening UBQLN-2 in French frontotemporal lobar degeneration and frontotemporal lobar degeneration-amyotrophic lateral sclerosis patients. Neurobiol Aging 34, 2078.e5–2078.e6 (2013).

59. Synofzik, M. et al. Screening in ALS and FTD patients reveals 3 novel UBQLN2 mutations outside the PXX domain and a pure FTD phenotype. Neurobiol Aging 33, 2949.e13–2949.e17 (2012).

60. Wu, Q. et al. Pathogenic Ubqln2 gains toxic properties to induce neuron death. Acta Neuropathol 129, 417–428 (2015).

61. Zhang, W., Huang, B., Gao, L. & Huang, C. Impaired 26s proteasome assembly precedes neuronal loss in mutant ubqln2 rats. Int J Mol Sci 22, (2021).

62. Chen, T., Huang, B., Shi, X., Gao, L. & Huang, C. Mutant UBQLN2P497H in motor neurons leads to ALS-like phenotypes and defective autophagy in rats. Acta Neuropathol Commun 6, 122 (2018).

63. Huang, B., Wu, Q., Zhou, H., Huang, C. & Xia, X. G. Increased Ubqln2 expression causes neuron death in transgenic rats. J Neurochem 285–293 (2016) doi:10.1111/jnc.13748.

64. Baglivo, M. et al. Early-onset of Frontotemporal Dementia and Amyotrophic Lateral Sclerosis in an Albanian Patient with a c.1319C>T Variant in the UBQLN2 Gene. Open Med J 7, 25–31 (2020).

65. Fadra, N. et al. Identification of skewed X chromosome inactivation using exome and transcriptome sequencing in patients with suspected rare genetic disease. BMC Genomics 25, 1–16 (2024).

66. Yang, Y., Jones, H. B., Dao, T. P. & Castañeda, C. A. Single Amino Acid Substitutions in Stickers, but Not Spacers, Substantially Alter UBQLN2 Phase Transitions and Dense Phase Material Properties. Journal of Physical Chemistry B 123, 3618–3629 (2019).

67. Kim, S. H. et al. Mutation-dependent aggregation and toxicity in a Drosophila model for UBQLN2-associated ALS. Hum Mol Genet 27, 322–337 (2018).

68. McCann, E. P. et al. Evidence for polygenic and oligogenic basis of Australian sporadic amyotrophic lateral sclerosis. J Med Genet 58, 87–95 (2021).

69. Matsumoto, A. et al. Unfavorable switching of skewed X chromosome inactivation leads to Menkes disease in a female infant. Sci Rep 14, 440 (2024).

70. Lupski, J. R., Garcia, C. A., Zoghbi, H. Y., Hoffman, E. P. & Fenwick, R. G. Discordance of muscular dystrophy in monozygotic female twins: Evidence supporting asymmetric splitting of the inner cell mass in a manifesting carrier of Duchenne dystrophy. Am J Med Genet 40, 354–364 (1991).

71. Dobrovolny, R. et al. Relationship between X-inactivation and clinical involvement in Fabry heterozygotes. Eleven novel mutations in the α-galactosidase A gene in the Czech and Slovak population. J Mol Med 83, 647–654 (2005).

72. Morrone, A. Fabry disease: molecular studies in Italian patients and X inactivation analysis in manifesting carriers. J Med Genet 40, 103e–1103 (2003).

73. Machiela, M. J. et al. Female chromosome X mosaicism is age-related and preferentially affects the inactivated X chromosome. Nat Commun 7, 11843 (2016).

74. Henden, L., et al. Short Tandem Repeat Expansions in Sporadic Amyotrophic Lateral Sclerosis and Frontotemporal Dementia. https://www.science.org (2023).

75. Van Rheenen, W. et al. Genome-wide association analyses identify new risk variants and the genetic architecture of amyotrophic lateral sclerosis. Nat Genet 48, 1043–1048 (2016).

76. Opie-Martin, S. et al. The SOD1-mediated ALS phenotype shows a decoupling between age of symptom onset and disease duration. Nat Commun 13, 1–9 (2022).

77. Keum, J. W. et al. The HTT CAG-Expansion Mutation Determines Age at Death but Not Disease Duration in Huntington Disease. Am J Hum Genet 98, 287–298 (2016).

