## Supplementary Figures for "A meta-analysis of genetic variant pathogenicity and sex differences in *UBQLN2*-linked amyotrophic lateral sclerosis and frontotemporal dementia"

Supplementary tables

Kyrah M. Thumbadoo^1,2^, Laura R. Nementzik^1,2^, Molly E.V. Swanson^1,2^, Birger V. Dieriks^2,3^, Michael Dragunow^2,4^, Richard L. M. Faull^2,3^, Maurice A. Curtis^2,3^, Ian P. Blair^5^, Garth A. Nicholson^5,6,7,8^, Kelly L. Williams^5^, Emma L. Scotter^1,2^

1 School of Biological Sciences, University of Auckland, Auckland 1010, New Zealand
2 Centre for Brain Research, University of Auckland, Auckland 1010, New Zealand
3 Department of Anatomy and Medical Imaging, University of Auckland, Auckland 1010, New Zealand
4 Department of Pharmacology and Clinical Pharmacology, University of Auckland, Auckland 1010, New Zealand
5 Macquarie University Centre for Motor Neuron Disease Research, Macquarie Medical School, Faculty of Medicine, Health and Human Sciences, Macquarie University, Sydney, New South Wales 2109, Australia
6 Northcott Neuroscience Laboratory, Australian and New Zealand Army Corps (ANZAC) Research Institute, Concord, New South Wales 2139, Australia
7 Faculty of Medicine, University of Sydney, Sydney, New South Wales 2050, Australia
8 Molecular Medicine Laboratory, Concord Repatriation General Hospital, Concord, New South Wales 2139, Australia

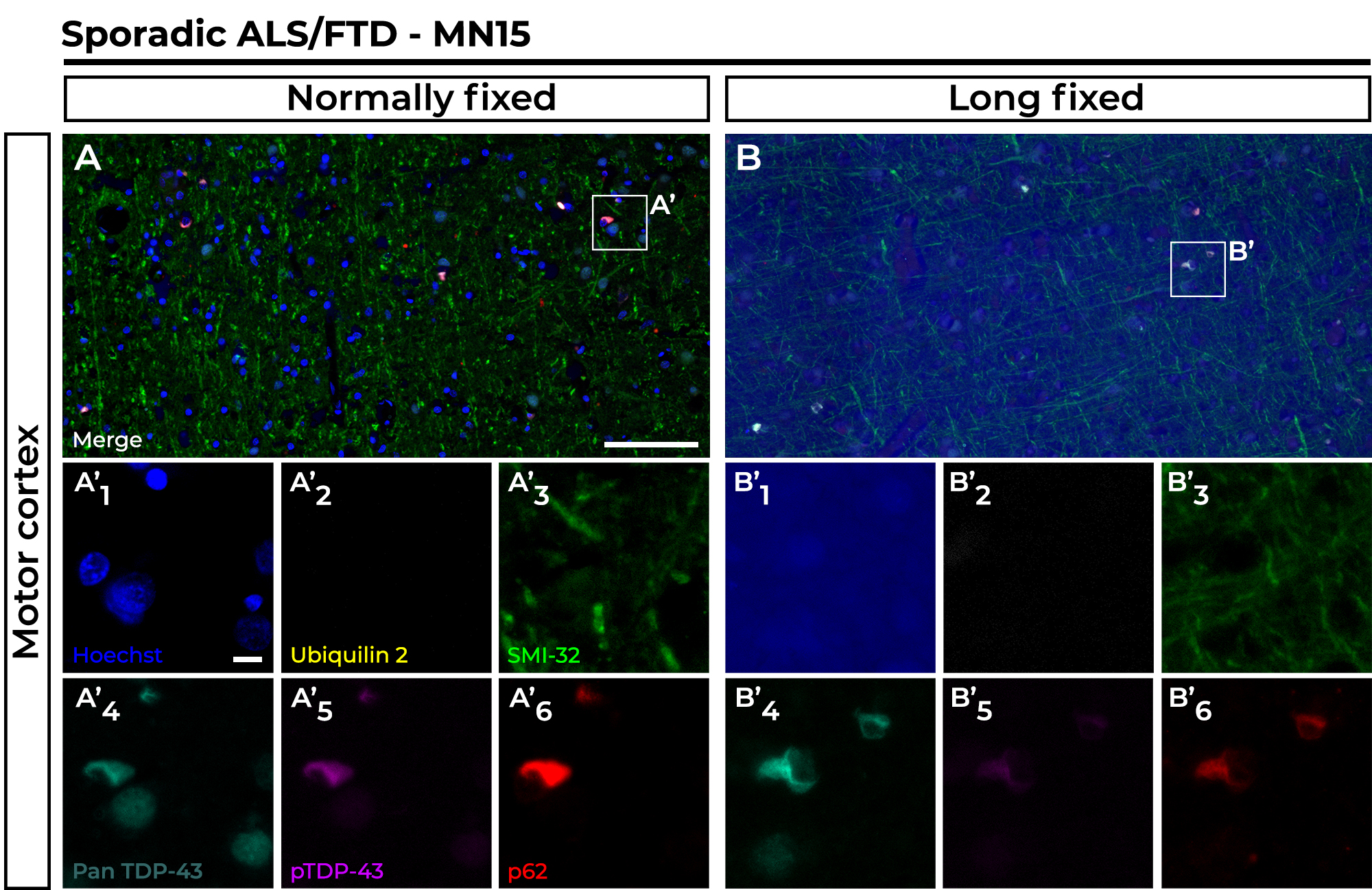

Supplementary Figure 1 Hoechst immunoreactivity is compromised but protein marker antigenicity is negligibly impacted by long-term fixation in formalin.

The motor cortex of case MN15 (sporadic ALS/FTD) from the right hemisphere (A) was processed and fixed normally and showed typical aggregate pathology of cytoplasmic phosphorylated TDP-43 (pTDP-43; BioLegend, #BL829901, 1:3000, RRID: AB_2564934) aggregates (A’_5_) that were co-labelled by p62 (Progen, #GP62-C, 1:500, RRID: AB_2687531) (A’_6_). The left hemisphere (B) of case MN15 showed the same aggregates (B’5 and B’6) despite being fixed in formalin for 10 years. Ubiquilin 2 pathology (A’_2_ and B’_2_) was not detected in either section, as expected. SMI-32 (Millipore, #AB5539, 1:500 RRID:AB_11212161), a neurofilament structural marker, showed no reduction in signal when long-fixed (A’_3_ and B’_3_). Of all markers tested, only Hoechst, a cell nuclei stain, was impaired with long fixation (A’_1_ and B’_1_). Scale bar in main images, 100 µm; 20 µm in zooms. Fluorescent immunohistochemistry was performed as per Methods in main article.

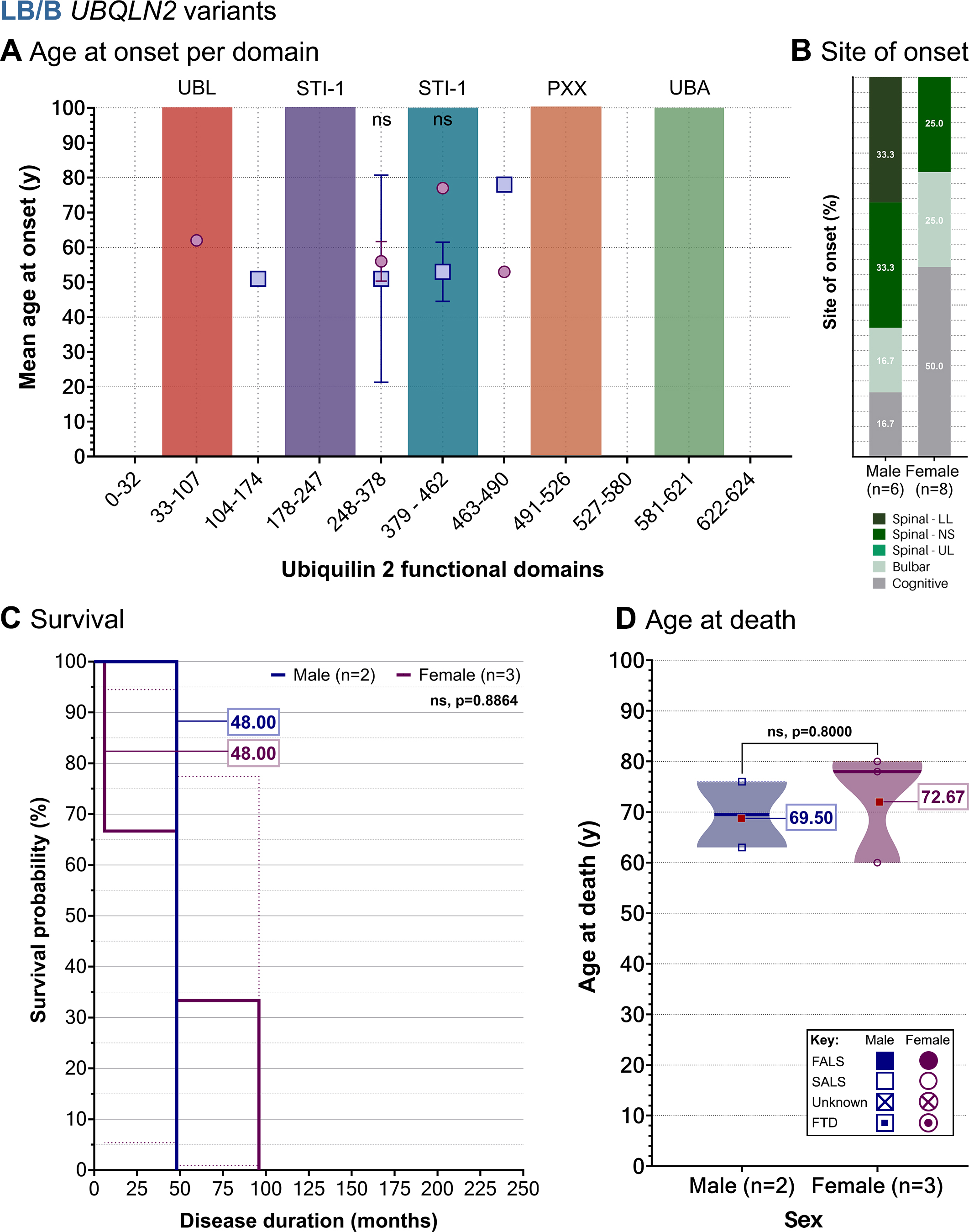

Supplementary Figure 2 For *UBQLN2* variants likely to be benign, no sex differences are observed in age at onset per *UBQLN2* functional domain, site of onset, survival, or age at death. Age at onset between males and females were compared across ubiquilin 2 domains in those harbouring likely benign and benign (A, LB/B) *UBQLN2* variants. Values indicate mean ± SD. Age at onset was compared using multiple *t*-tests corrected for multiple comparisons by Holm-Šídák method. (B) Site of onset distributions were not found to be significantly different due to absence of for some onset sites per sex group. (C) Survival information was available for two males and three females, showing the same median survival time of 48 months. (D) Age at death was available for three females and three males, showing a non-significant difference of 3.17 years. Abbreviations: FALS, familial ALS; FTD, frontotemporal dementia; ns, not significant; spinal – LL, spinal – lower limb; spinal – NS, spinal – not specified; spinal – UL, spinal – upper limb; SALS, sporadic ALS.
